# What variables can better predict the number of infections and deaths worldwide by SARS-CoV-2? Variation through time

**DOI:** 10.1101/2020.06.04.20122176

**Authors:** J. G. García de Alcañíz, J. Romero-López, R. P. Martínez-Esteban, V. López-Rodas, E. Costas

## Abstract

Using data from 50 very different countries (which represent nearly 70% of world’s population) and by means of a regression analysis, we studied the predictive power of different variables (mobility, air pollution, health & research, economic and social & geographic indicators) over the number of infected and dead by SARS-CoV-2. We also studied if the predictive power of these variables changed during a 4 months period (March, April, May and June). We approached data in two different ways, cumulative data and non-cumulative data.

The number of deaths by Covid-19 can always be predicted with great accuracy from the number of infected, regardless of the characteristics of the country.

Inbound tourism emerged as the variable that best predicts the number of infected (and, consequently, the number of deaths) happening in the different countries. Electricity consumption and air pollution of a country (CO_2_ emissions, nitrous oxide and methane) are also capable of predicting, with great precision, the number of infections and deaths from Covid-19. Characteristics such as the area and population of a country can also predict, although to a lesser extent, the number of infected and dead. All predictive variables remained significant through time.

In contrast, a series of variables, which in principle would seem to have a greater influence on the evolution of Covid-19 (hospital bed density, Physicians per 1000 people, Researches in R & D, urban population…), turned out to have very little - or none- predictive power.

Our results explain why countries that opted for travel restrictions and social withdrawal policies at a very early stage of the pandemic outbreak, obtained better results. Preventive policies proved to be the key, rather than having large hospital and medical resources.

## INTRODUCTION

COVID-19 is a global pandemic caused by SARS-CoV-2 virus^1,2^. The virus rapidly spread and is happening at the same time all around the world, to different people living in different countries (with different mobility, pollution, health, economic, geographic and social characteristics). Infection and death counts vary widely not only from country to country but also through time. In some countries the virus is in the remission phase, with chances of new outbreaks, while in others is yet uncontrolled.

The main feature of COVID-19 pandemic is the lack of knowledge, SARS-CoV-2 is a new coronavirus and little is known yet, despite the effort of scientists around the world. Under this setting, regression analysis is a very useful tool to find out which variables can predict number of infected and dead, and which variables cannot. Consequently, numerous studies have been performed since the outbreak of the pandemic using regression analysis ^3–7^, most of them focus on the clinical aspects of the pandemic and its implications on health care settings ^5,8–13^, such as cardiac injury, kidney disease, respiratory conditions, diabetes, comorbidities ^8,9,14–19^ gender and underlying comorbidities ^20^, severe COVID-19 condition and risk factors ^21–24^, number of CD8 +T cells as risk factors for the duration of SARS-CoV-2 viral positivity ^25 26^, clinical symptoms as smell loss ^27^, smoking habits ^28^. And not as many, on other aspects like looking into, for example, temperature as an affecting parameter to transmission rates^3,29–31^. Some have used GIS modelling to understand what is happening with SARS CoV-2 in this pandemic ^32^. Some others try to unveil the relevance of temperature, humidity, climate, air quality, socioeconomic factors, demographics…^3,4,38–40,5,30,31,33–37^.

The relationship of COVID-19 with socioeconomic variables has also been investigated. For example, Mollalo et al. (2020) have performed a study in the U.S.A. compiling 35 environmental, socioeconomic, topographic and demographic variables and create a geodatabase to explain the spatial variability of the COVID incidence ^37^. GIS-based spatial modelling studies have also been done.^32^ Guha et al (2020) studied community and socioeconomic characteristics of people in the United States and its possible associations with COVID-19 cases and deaths^6^, Whittle et al (2020) studied socioeconomic predictors across neighbourhoods in New York ^7^, Pirouz et al (2020) mixed climate and urban parameters for three regions in Italy ^4^

In this study we used a regression analysis between the number of COVID-19 cases and the number of deaths, the dependent variables; and a series of independent variables, such as Mobility, Air Pollution, Health & Research, Economic, Geographical and Social Indicators, to estimate the predictive value of those independent variables and if it changed with time. We also analysed how the predictive value of these variables changed through May and June but using non-cumulative data.

Interestingly, variables related to mobility and air pollution better explain number of infected and dead from COVID-19 than other variables that estimate countries health system’s quality or economic strength.

## MATERIALS AND METHODS

We carried out a regression analysis, trying to unveil which characteristics make a country more vulnerable to Covid19.

The chosen dependent variables correspond to two of the most significant and worrisome characteristics of any outbreak: total number of infected people and total death toll. Data was collected from the World Health Organisation (WHO) Coronavirus Disease (COVID-19) Dashboard^41^ (retrieved from https://covid19.who.int the 21^st^ of: March, April, May and June).

This study focuses on the predictive ability -or not- of the chosen variables, regarding infected and death counts. Calculations were performed using data in two different ways: cumulative count and non-cumulative count. Cumulative count data implies that figures from one period include previous ones. Non-cumulative count represent exclusively figures from the period of time studied (i.e. May period starts on the 21^st^ of April and ends on the 21^st^ May; June period starts on the 21^st^ of May and ends on the 21^st^ of June).

We first analysed how number of infected predicts number of deaths and its evolution through time. If there was a significant improvement in treatment protocols, its predictive value should change.

The predictive variables were selected searching for relations that may explain the differences within countries. Mobility easiness also implies an easy way to spread pathogens, we selected Inbound Tourism as an international mobility indicator. One Health speaks about Human Health, Animal Health and Environmental Health: we selected Air Pollution indicators such as CO_2_, NO_2_ and methane emissions, as they may show an influence on the disease incidence. Health indicators (e.g. Hospital Bed Density -beds per 1000- or Physicians per 1000 people, Researches in R & D). Economic indicators as different outcomes could be influenced by the countries’ wealth. Social and geographic indicators to study any pattern that explained countries differences (e.g. population, surface area, urban population…).

All data was obtained via internet repositories open to public access, such as The World Health Organization, The Central Intelligence Agency or The World Bank. The complete list of variables, as well as the link to each data set is presented in Table 1.

**Table 1.** List of predictive variables and data source

| Predictive Variable | Source |
| --- | --- |
| *International Mobility Indicators* |  |
| Inbound Tourism | https://www.unwto.org/es/country-profile-inbound-tourism |
| *Air Pollution Indicators* |  |
| Electricity Consumption | https://www.cia.gov/library/publications/the-world-factbook/fields/253rank.html |
| Carbon dioxide emissions from consumption of energy | https://www.cia.gov/library/publications/the-world-factbook/fields/274rank.html |
| Nitrous oxide emissions (thousand metric tons of CO<sub>2</sub> equivalent) | https://data.worldbank.org/indicator/EN.ATM.NOXE.KT.CE |
| Methane emissions (kT of CO<sub>2</sub> equivalent) | https://data.worldbank.org/indicator/EN.ATM.METH.KT.CE |
| *Health and Research Indicators* |  |
| Hospital Bed Density (beds per 1000 people) | https://www.cia.gov/library/publications/the-world-factbook/fields/360.html |
| Physicians (per 1,000 people) | https://data.worldbank.org/indicator/SH.MED.PHYS.ZS |
| Researches in R & D (per million people) | https://data.worldbank.org/indicator/SP.POP.SCIE.RD.P6 |
| *Economic Indicators* |  |
| Unemployment rate | https://www.cia.gov/library/publications/resources/the-world-factbook/fields/220rank.html |
| Inflation rate (consumer prices) | https://www.cia.gov/library/publications/resources/the-world-factbook/fields/229rank.html |
| Government expenditure on education, total (% of GDP) | https://data.worldbank.org/indicator/SE.XPD.TOTL.GD.ZS |
| Military expenditures | https://www.cia.gov/library/publications/resources/the-world-factbook/fields/330rank.html |

| Social and Demographic Indicators |  |
| --- | --- |
| Population: | https://www.cia.gov/library/publications/resources/the-world-factbook/fields/335rank.html |
| Area: | https://www.cia.gov/library/publications/resources/the-world-factbook/fields/279rank.html |
| Urban Population | https://data.worldbank.org/indicator/SP.URB.TOTL.IN.ZS?view=chart |
| Median age | https://www.cia.gov/library/publications/resources/the-world-factbook/fields/343rank.html |
| Population ages 65 and above | https://data.worldbank.org/indicator/SP.POP.65UP.TO.ZS |
| Proportion of seats held by women in national parliaments | https://data.worldbank.org/indicator/SG.GEN.PARL.ZS |
| Population Growth rate | https://www.cia.gov/library/publications/resources/the-world-factbook/fields/344rank.html |
| Infant Mortality Rate | https://www.cia.gov/library/publications/resources/the-world-factbook/fields/354rank.html |
| Life expectancy at birth | https://www.cia.gov/library/publications/resources/the-world-factbook/fields/355rank.html |
| Obesity - Adult prevalence rate | https://www.cia.gov/library/publications/resources/the-world-factbook/fields/367rank.html |

The territories for this study were the first 50 countries (which represent nearly 70% of world’s population) with more inbound tourism (last data available from 2018), according to the World Tourism Organization’s webpage^42^ (https://www.unwto.org/country-profile-inbound-tourism). In these countries, very different to each other but with lots of people entering from everywhere, the likelihood of a quick SARS-CoV-2 arrival was very high.

Regression analysis allows to establish a relation between two variables, a dependent variable y_i_ and an independent variable x_i_. This linear regression follows the model:

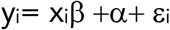

and was performed using the statistic package StatPlus^:mac^ (AnalystSoft Inc.)

## RESULTS & DISCUSSION

Before any further analysis we studied the relation between the number of dead by SARS-COV-2 (y_i_= dependent variable) and the number of infected people (x_i_= predictive variable). Among all the studied parameters, these had the highest significant regression (p<0.00001) consistently through the period studied (Cumulative March: R^2^=0.80; C. April: R^2^=0.76; C. May: R^2^=0.99; C. June: R^2^=0.85).

The coefficient of determination (R^2^) is the proportion of the variance in the dependent variable that is predictable from the independent variable (e.g. in C. May 99% of the deaths were explained by the number of infected people).

Furthermore, when non-cumulative data was used results were as significant and surprising as before (Non-Cumulative May: R^2^=0.85; N-C. June: R^2^=0.85). We were expecting some change as health care systems learnt how to fight the virus. This may appear as an obvious conclusion, but it gains relevance when you consider that other parameters studied (i.e. health and research indicators, country richness…) had no significance, this is in line with Walach et al (2020) findings, that none of the variables related to Health or other population parameters were predictive^43^. No matter what country you choose (rich, poor, with a good or not so good health system, etc.) the number of infected people is going to rule the number of deaths. This drives the idea that what really matters to fight COVID-19 pandemic is to prevent infections among their citizens rather than better health care systems or richness or development. This may be the reason behind why poorer countries with less developed health care systems did so well fighting this pandemic outbreak (e.g. Viet Nam).

We present variables divided into clearly predictive and not predictive. As interesting is to know which ones can predict as it is to know the ones that do not predict at all.

### 1. Variables that are clearly predictive of the covid-19 pandemic (statistically significant linear regression p < 0.05)

Inbound tourism was the second variable with greater significance. It remains significant with the number of infected people during the cumulative four-month period studied (C. March: p<0.00001, R^2^=0.34; C. April: p<0.00001, R^2^=0.42; C. May: p<0.00006, R^2^=0.29; C. June: p<0.0003 R^2^=0,17) and with non-cumulative data (N-C May: p<0.005, R^2^=0.15; N-C June: p<0.0003 R^2^=0.17). Our results are concordant with the results obtained by Aldibasi et al (2020)^44^. Daon et al (2020) suggest that an additional prominent reason for the especially wide and rapid spread of COVID-19 is likely to be the current high prevalence of international travel^45^. According to Papadopoulus et al. (2020) early international travel restrictions may help control the pandemic^46^, and Walch et al (2020) found that border closure had the potential of preventing cases^43^. For Chinazzi et al. (2020) most of the imported cases outside China have likely originated from air travel^47^ and Linka et al. suggest that the spread of COVID-19 in Europe closely followed air travel patterns and that the severe travel restrictions implemented there resulted in substantial decreases in the disease’s spread^48^. Kubota et al (2020) found that relative frequency of foreign visitors per population was positively correlated^49^ and Coelho et al. (2020) emphasized the role of the air transportation network in this epidemic^50^. Gross et al studying spatial dynamics of the COVID-19 in China found a strong correlation between the number of infected individuals in each province studied and the population migration from Hubei to those provinces^51^ with a slight decay along time, which agrees with our results although remaining highly significant. Tuli et al (2020) show in their model that countries with higher air passenger traffic have higher likelihood of getting higher level of infections^52^. *Lockdown-type measures had the largest effect on limiting viral transmission, followed by complete travel ban*^53^

Inbound tourism also has a significant association with the other dependent variable, number of dead people and stays significant with both cumulative (C. March: p<0.0003, R^2^=0.24; C. April: p<0.00001, R^2^=0.63; C. May: p<0.00001, R^2^=0.46; C. June: p=0.00001, R^2^=0.35) and non-cumulative data (N-C May: p<0.0001, R^2^=0.27; N-C June: p<0.00001, R^2^=0.35).

This means that the number of people arriving to a country is linked to number of infected or dead by SARS-CoV-2. This is concordant with the results obtained by Anzai et al.^54^. Merler et al. conducted a study to see the effect of travel restrictions on the spread of the virus, and they found that the travel quarantine of Wuhan delayed the overall epidemic progression by only 3 to 5 days in mainland China but had a more marked effect on the international scale, where case importations were reduced by nearly 80% until mid-February ^55^.

Other results that we estimated relevant was the strong regression found between infected and dead by COVID-19 and independent variables that talk about air pollution.

Electricity consumption had high statistical significance with both dependent variables, number of infected people and number of deaths, and were statistically significant with cumulative data and non-cumulative data. Other environmental indicators (CO_2_ emissions, NO_2_ emissions and methane emissions) also showed high predictive power since they had statistical significance (with p<0.001 values) Countries with more emissions and more energy usage had higher counts of infected or dead people (Results are shown on Table 2 and Table 3).

**Table 2:** Cumulative Data. Variables that are clearly predictive of the covid-19 pandemic (statistically significant linear regression p < 0.05)

| **I. CUMULATIVE DATA. VARIABLES THAT ARE CLEARLY PREDICTIVE OF THE COVID-19 PANDEMIC**<br>(statistically significant linear regression $p < 0.05$)<br>21<sup>st</sup> of MARCH 2020 | | | | | | | |
| --- | --- | --- | --- | --- | --- | --- | --- |
| dependent variable<br>(y) | predictive variable<br>(x) | F value | d. f. regression<br>residue | P | R | R<sup>2</sup> | $\alpha$ and $\beta$ coefficients |
| **Dead vs. infected covid-19** |  |  |  |  |  |  |  |
| dead by covid-19 | infected by covid-19 | 194.20 | 1<br>48 | <0.00001 *** | 0.90 | 0.80 | $\alpha=-44.44$<br>$\beta=0.0531$ |
| **International Mobility Indicators** |  |  |  |  |  |  |  |
| infected by covid-19 | Inbound tourism arrivals (in millions) | 24.73 | 1<br>48 | 0.00001 *** | 0.58 | 0.34 | $\alpha=-3605.16$<br>$\beta=404.06$ |
| dead by covid-19 | Inbound tourism arrivals (in millions) | 15.23 | 1<br>48 | 0.0003 *** | 0.49 | 0.24 | $\alpha=-206$<br>$\beta=20.15$ |
| **Air Pollution Indicators** |  |  |  |  |  |  |  |
| infected by covid-19 | electricity consumption | 10.76 | 1<br>48 | <0.00001 *** | 0.69 | 0.47 | $\alpha=1407.39$<br>$\beta<0.00001$ |
| dead by covid-19 | electricity consumption | 10.35 | 1<br>43 | 0.0023 *** | 0.42 | 0.17 | $\alpha=101.56$<br>$\beta<0.00001$ |
| infected by covid-19 | Carbon dioxide emissions | 55.34 | 1<br>48 | <0.00001 *** | 0.73 | 0.54 | $\alpha=1604.23$<br>$\beta=0.00001$ |
| dead by covid-19 | Carbon dioxide emissions | 12.83 | 1<br>48 | 0.0008*** | 0.46 | 0.21 | $\alpha=104.38$<br>$\beta<0.00001$ |
| infected by covid-19 | Nitrous oxide emissions | 29.32 | 1<br>48 | <0.00001 *** | 0.65 | 0.43 | $\alpha=26250$<br>$\beta=0.4545$ |
| dead by covid-19 | Nitrous oxide emissions | 8.59 | 1<br>39 | 0.0056** | 0.43 | 0.18 | $\alpha=125.52$<br>$\beta=0.0939$ |
| infected by covid-19 | Methane emissions | 28,62 | 1<br>40 | <0.00001 *** | 0.65 | 0.42 | $\alpha=35022$<br>$\beta=0.1012$ |
| dead by covid-19 | Methane emissions | 8.62 | 1<br>39 | 0.0055** | 0.42 | 0.18 | $\alpha=135$<br>$\beta=0.00133$ |
| **Social and Geographic Indicators** |  |  |  |  |  |  |  |
| infected by covid-19 | Population | 10.90 | 1<br>48 | 0.00007*** | 0.53 | 0.28 | $\alpha=2520$<br>$\beta=0.00003$ |
| dead by covid-19 | Population | 6.48 | 1<br>48 | 0.014* | 0.34 | 0.12 | $\alpha=134$<br>$\beta<00001$ |
| infected by covid-19 | Area | 7,70 | 1<br>48 | 0.0049 *** | 0.39 | 0.1153 | $\alpha=2626$<br>$\beta=0.0015$ |
| dead by covid-19 | Area | 8.51 | 1<br>48 | 0.0053 ** | 0.39 | 0.15 | $\alpha=78.14$<br>$\beta=0.00009$ |

| **I. CUMULATIVE DATA. VARIABLES THAT ARE CLEARLY PREDICTIVE OF THE COVID-19 PANDEMIC**<br>(statistically significant linear regression $p < 0.05$)<br>21<sup>st</sup> of APRIL 2020 | | | | | | | |
| --- | --- | --- | --- | --- | --- | --- | --- |
| dependent variable<br>(y) | predictive variable<br>(x) | F value | d. f. regression<br>residue | P | R | R<sup>2</sup> | $\alpha$ and $\beta$ coefficients |
| **Dead vs. infected covid-19** |  |  |  |  |  |  |  |
| dead by covid-19 | infected by covid-19 | 136.20 | 1<br>43 | <0.00001 *** | 0.87 | 0.76 | $\alpha=-35423$<br>$\beta=3580$ |
| **International Mobility Indicators** |  |  |  |  |  |  |  |
| infected by covid-19 | Inbound tourism arrivals (in millions) | 31.08 | 1<br>43 | 0.00001 *** | 0.65 | 0.42 | $\alpha=786$<br>$\beta=0.057$ |
| dead by covid-19 | Inbound tourism arrivals (in millions) | 73.49 | 1<br>43 | 0.00001 *** | 0.79 | 0.63 | $\alpha=-3136$<br>$\beta=285$ |
| **Air Pollution Indicators** |  |  |  |  |  |  |  |
| infected by covid-19 | electricity consumption | 20.76 | 1<br>43 | 0.00001 *** | 0.57 | 0.33 | $\alpha=18774$<br>$\beta<0.00001$ |
| dead by covid-19 | electricity consumption | 8.32 | 1<br>43 | 0.006 *** | 0.40 | 0.16 | $\alpha=2162$<br>$\beta<0.00001$ |
| infected by covid-19 | Carbon dioxide emissions | 8.71 | 1<br>43 | 0.005 *** | 0.41 | 0.17 | $\alpha=28961$<br>$\beta=0.00003$ |
| dead by covid-19 | Carbon dioxide emissions | 3.37 | 1<br>43 | 0.0735 | 0.27 | 0.07 | $\alpha=2690$<br>$\beta<0.00001$ |
| infected by covid-19 | Nitrous oxide emissions | 7.99 | 1<br>43 | 0.007 ** | 0.40 | 0.16 | $\alpha=26250$<br>$\beta=0.4545$ |
| dead by covid-19 | Nitrous oxide emissions | 3.24 | 1<br>43 | 0.08 n.s. | 0.26 | 0.07 | $\alpha=2558$<br>$\beta=0.01974$ |
| infected by covid-19 | Methane emissions | 2.85 | 1<br>43 | 0.10 n.s. | 0.25 | 0.06 | $\alpha=35022$<br>$\beta=0.1012$ |
| dead by covid-19 | Methane emissions | 0.90 | 1<br>43 | 0.34 n.s. | 0.14 | 0.02 | $\alpha=3017$<br>$\beta=0.00377$ |
| **Social and Geographic Indicators** |  |  |  |  |  |  |  |
| infected by covid-19 | Population | 1.09 | 1<br>43 | 0.30 n.s. | 0.157 | 0.024 | $\alpha=40190$<br>$\beta=0.00007$ |
| dead by covid-19 | Population | 0.32 | 1<br>43 | 0.57 n.s. | 0.086 | 0.007 | $\alpha=3221$<br>$\beta<0.00001$ |
| infected by covid-19 | Area | 7,45 | 1<br>43 | 0.009 ** | 0.384 | 0.147 | $\alpha=22664$<br>$\beta=0.01185$ |
| dead by covid-19 | Area | 4.31 | 1<br>43 | 0.04 * | 0.30 | 0.09 | $\alpha=2211$<br>$\beta=0.00006$ |

| I. CUMULATIVE DATA. VARIABLES THAT ARE CLEARLY PREDICTIVE OF THE COVID-19 PANDEMIC<br>(statistically significant linear regression $p < 0.05$)<br>21<sup>st</sup> of MAY 2020 | | | | | | | |
| --- | --- | --- | --- | --- | --- | --- | --- |
| dependent variable<br>(y) | predictive variable<br>(x) | F value | d. f. regression<br>residue | P | R | R<sup>2</sup> | $\alpha$ and $\beta$ coefficients |
| **Dead vs. infected covid-19** |  |  |  |  |  |  |  |
| dead by<br>covid-19 | infected by covid-19 | 4912108 | 1<br>48 | <0.00001 *** | 0.99 | 0.99 | $\alpha=-0.22617$<br>$\beta=0.02910$ |
| **International Mobility Indicators** |  |  |  |  |  |  |  |
| infected by covid-19 | Inbound tourism<br>arrivals (in millions) | 19.52 | 1<br>48 | 0.00006 *** | 0.54 | 0.29 | $\alpha=-43061$<br>$\beta=05702$ |
| dead by<br>covid-19 | Inbound tourism<br>arrivals (in millions) | 40.22 | 1<br>48 | <0.00001 *** | 0.67 | 0.46 | $\alpha=-4900$<br>$\beta=485$ |
| **Air Pollution Indicators** |  |  |  |  |  |  |  |
| infected by covid-19 | electricity<br>consumption | 22.47 | 1<br>48 | 0.00002 *** | 0.56 | 0.32 | $\alpha=34430$<br>$\beta<0.00001$ |
| dead by<br>covid-19 | electricity<br>consumption | 14.00 | 1<br>48 | 0.00049 *** | 0.47 | 0.23 | $\alpha=3116$<br>$\beta<0.00001$ |
| infected by covid-19 | Carbon dioxide<br>emissions | 9.20 | 1<br>48 | 0.004 ** | 0.40 | 0.16 | $\alpha=53388$<br>$\beta=0.00005$ |
| dead by<br>covid-19 | Carbon dioxide<br>emissions | 5.25 | 1<br>48 | 0.026* | 0.31 | 0.10 | $\alpha=4383$<br>$\beta<0.00001$ |
| infected by covid-19 | Nitrous oxide<br>emissions | 3.31 | 1<br>39 | 0.08 n.s. | 0.28 | 0.08 | $\alpha=48735$<br>$\beta=0.2236$ |
| dead by<br>covid-19 | Nitrous oxide<br>emissions | 0.51 | 1<br>39 | 0.48 n.s. | 0.11 | 0.013 | $\alpha=3930$<br>$\beta=0.009$ |
| infected by covid-19 | Methane emissions | 4.91 | 1<br>39 | 0.03* | 0.33 | 0.11 | $\alpha=47540$<br>$\beta=0.09$ |
| dead by<br>covid-19 | Methane emissions | 0.21 | 1<br>39 | 0.65 n.s. | 0.07 | 0.005 | $\alpha=4083$<br>$\beta=0.0019$ |
| **Social and Geographic Indicators** |  |  |  |  |  |  |  |
| infected by covid-19 | Population | 1.62 | 1<br>48 | 0.20 n.s. | 0.18 | 0.032 | $\alpha=70907$<br>$\beta=0.00015$ |
| dead by<br>covid-19 | Population | 0.68 | 1<br>48 | 0.41 n.s. | 0.12 | 0.014 | $\alpha=5450$<br>$\beta<0.00001$ |
| infected by covid-19 | Area | 15,38 | 1<br>48 | 0.0002 *** | 0.49 | 0.243 | $\alpha=29559$<br>$\beta=0.02954$ |
| dead by<br>covid-19 | Area | 7.86 | 1<br>48 | 0.007 ** | 0.37 | 0.14 | $\alpha=3202$<br>$\beta=0.0015$ |

| **I. CUMULATIVE DATA. VARIABLES THAT ARE CLEARLY PREDICTIVE OF THE COVID-19 PANDEMIC**<br>(statistically significant linear regression $p < 0.05$)<br>21<sup>st</sup> of JUNE 2020 | | | | | | | |
| --- | --- | --- | --- | --- | --- | --- | --- |
| dependent variable<br>(y) | predictive variable<br>(x) | F value | d. f. regression<br>residue | P | R | R<sup>2</sup> | $\alpha$ and $\beta$ coefficients |
| **Dead vs. infected covid-19** |  |  |  |  |  |  |  |
| dead by covid-19 | infected by covid-19 | 269.83 | 1<br>48 | <0.00001 *** | 0.92 | 0.85 | $\alpha=977.02$<br>$\beta=0.05189$ |
| **International Mobility Indicators** |  |  |  |  |  |  |  |
| infected by covid-19 | Inbound tourism arrivals (in millions) | 965 | 1<br>48 | 0.0003 *** | 0.41 | 0.17 | $\alpha=-10030$<br>$\beta=6857$ |
| dead by covid-19 | Inbound tourism arrivals (in millions) | 25.49 | 1<br>48 | =0.00001 *** | 0.59 | 0.35 | $\alpha=-4108$<br>$\beta=556$ |
| **Air Pollution Indicators** |  |  |  |  |  |  |  |
| infected by covid-19 | electricity consumption | 18.57 | 1<br>48 | 0.00008 *** | 0.53 | 0.28 | $\alpha=68670$<br>$\beta<0.00001$ |
| dead by covid-19 | electricity consumption | 14.43 | 1<br>48 | 0.00041 *** | 0.48 | 0.23 | $\alpha=4588$<br>$\beta<0.00001$ |
| infected by covid-19 | Carbon dioxide emissions | 7.77 | 1<br>48 | 0.0075 ** | 0.37 | 0.14 | $\alpha=96950$<br>$\beta=0.00007$ |
| dead by covid-19 | Carbon dioxide emissions | 5.41 | 1<br>48 | 0.024* | 0.32 | 0.10 | $\alpha=6196$<br>$\beta<0.00001$ |
| infected by covid-19 | Nitrous oxide emissions | 5.49 | 1<br>39 | 0.024 * | 0.35 | 0.12 | $\alpha=81526$<br>$\beta=0.0683$ |
| dead by covid-19 | Nitrous oxide emissions | 2.42 | 1<br>39 | 0.13 n.s. | 0.24 | 0.06 | $\alpha=5218$<br>$\beta=0.0262$ |
| infected by covid-19 | Methane emissions | 5.74 | 1<br>39 | 0.021* | 0.36 | 0.13 | $\alpha=82736$<br>$\beta=0.2293$ |
| dead by covid-19 | Methane emissions | 1.37 | 1<br>39 | 0.25 n.s. | 0.18 | 0.003 | $\alpha=5544$<br>$\beta=0.0067$ |
| **Social and Geographic Indicators** |  |  |  |  |  |  |  |
| infected by covid-19 | Population | 2.92 | 1<br>48 | 0.09 n.s. | 0.24 | 0.06 | $\alpha=112655$<br>$\beta=0.00031$ |
| dead by covid-19 | Population | 1.45 | 1<br>48 | 0.23 n.s. | 0.17 | 0.03 | $\alpha=7215$<br>$\beta=0.00001$ |
| infected by covid-19 | Area | 20.40 | 1<br>48 | 0.00004 *** | 0.55 | 0.30 | $\alpha=45752$<br>$\beta=0.5175$ |
| dead by covid-19 | Area | 11.24 | 1<br>48 | 0.0015 ** | 0.43 | 0.19 | $\alpha=4051$<br>$\beta=0.00232$ |

**Table 3:** Non-Cumulative Data. Variables that are clearly predictive of the covid-19 pandemic (statistically significant linear regression p < 0.05)

| **II. NON-CUMULATIVE DATA. VARIABLES THAT ARE CLEARLY PREDICTIVE OF THE COVID-19 PANDEMIC**<br>(statistically significant linear regression $p < 0.05$)<br>Non-Cumulative MAY | | | | | | | |
| --- | --- | --- | --- | --- | --- | --- | --- |
| dependent variable<br>(y) | predictive variable<br>(x) | F value | d. f. regression<br>residue | P | R | R<sup>2</sup> | $\alpha$ and $\beta$ coefficients |
| **Dead vs. infected Covid-19** |  |  |  |  |  |  |  |
| dead by covid-19 | infected by covid-19 | 277 | 1<br>48 | <0.000001*** | 0.92 | 0.85 | $\alpha=161$<br>$\beta=0.067$ |
| **International Mobility Indicators** |  |  |  |  |  |  |  |
| infected by covid-19 | Inbound tourism arrivals (in millions) | 8.55 | 1<br>48 | 0.0052 *** | 0.39 | 0.15 | $\alpha=7108$<br>$\beta=2155$ |
| dead by covid-19 | Inbound tourism arrivals (in millions) | 17.61 | 1<br>48 | 0.0001 *** | 0.52 | 0.27 | $\alpha=-3136$<br>$\beta=285$ |
| **Air Pollution Indicators** |  |  |  |  |  |  |  |
| infected by covid-19 | electricity consumption | 17.77 | 1<br>48 | 0.0001 *** | 0.52 | 0.27 | $\alpha=16750$<br>$\beta=0.00001$ |
| dead by covid-19 | electricity consumption | 15.16 | 1<br>48 | 0.0003 *** | 0.49 | 0.24 | $\alpha=1274$<br>$\beta=0.00001$ |
| infected by covid-19 | Carbon dioxide emissions | 7.26 | 1<br>48 | 0.009 *** | 0.36 | 0.13 | $\alpha=26171$<br>$\beta=0.00002$ |
| dead by covid-19 | Carbon dioxide emissions | 5.39 | 1<br>48 | 0.02* | 0.32 | 0.10 | $\alpha=1967$<br>$\beta=0.00001$ |
| infected by covid-19 | Nitrous oxide emissions | 2.42 | 1<br>39 | 0.13 n.s. | 0.24 | 0.06 | $\alpha=21943$<br>$\beta=0.1315$ |
| dead by covid-19 | Nitrous oxide emissions | 1.51 | 1<br>39 | 0.22 n. s. | 0.19 | 0.037 | $\alpha=1512$<br>$\beta=0.00594$ |
| infected by covid-19 | Methane emissions | 4,59 | 1<br>39 | 0.04* | 0.32 | 0.10 | $\alpha=20458$<br>$\beta=0.059$ |
| dead by covid-19 | Methane emissions | 0.69 | 1<br>39 | 0.41 n. s. | 0.13 | 0.018 | $\alpha=1604$<br>$\beta=0.00138$ |
| **Social & Geographic Indicators** |  |  |  |  |  |  |  |
| infected by covid-19 | Population | 1.72 | 1<br>48 | 0.19 n. s. | 0.19 | 0.034 | $\alpha=-33384$<br>$\beta=0.00008$ |
| dead by covid-19 | Population | 0.80 | 1<br>48 | 0.37 n. s. | 0.128 | 0.016 | $\alpha=2542$<br>$\beta<0.00001$ |
| infected by covid-19 | Area | 21,94 | 1<br>48 | 0.00002 *** | 0.56 | 0.31 | $\alpha=7912$<br>$\beta=0.0175$ |
| dead by covid-19 | Area | 8.81 | 1<br>48 | 0.004 ** | 0.39 | 0.16 | $\alpha=1236$<br>$\beta=0.0009$ |

| **II. NON-CUMULATIVE DATA. VARIABLES THAT ARE CLEARLY PREDICTIVE OF THE COVID-19 PANDEMIC**<br>(statistically significant linear regression $p < 0.05$)<br>**Non-Cumulative JUNE** | | | | | | | |
| --- | --- | --- | --- | --- | --- | --- | --- |
| dependent variable<br>(y) | predictive variable<br>(x) | F value | d. f. regression<br>residue | P | R | R<sup>2</sup> | $\alpha$ and $\beta$ coefficients |
| **Dead vs. infected Covid-19** |  |  |  |  |  |  |  |
| dead by covid-19 | infected by covid-19 | 269 | 1<br>48 | <0.000001*** | 0.92 | 0.85 | $\alpha=6197$<br>$\beta=16.35$ |
| **International Mobility Indicators** |  |  |  |  |  |  |  |
| infected by covid-19 | Inbound tourism arrivals (in millions) | 9.65 | 1<br>48 | 0.0003 *** | 0.41 | 0.17 | $\alpha=-10030$<br>$\beta=6858$ |
| dead by covid-19 | Inbound tourism arrivals (in millions) | 25.49 | 1<br>48 | 0.00001 *** | 0.59 | 0.35 | $\alpha=-4108$<br>$\beta=556$ |
| **Air Pollution Indicators** |  |  |  |  |  |  |  |
| infected by covid-19 | electricity consumption | 18.57 | 1<br>48 | 0.0008 *** | 0.53 | 0.27 | $\alpha=66670$<br>$\beta<0.00001$ |
| dead by covid-19 | electricity consumption | 14.43 | 1<br>48 | 0.0004 *** | 0.48 | 0.23 | $\alpha=4588$<br>$\beta<0.00001$ |
| infected by covid-19 | Carbon dioxide emissions | 7.76 | 1<br>48 | 0.007 *** | 0.37 | 0.14 | $\alpha=96950$<br>$\beta=0.00007$ |
| dead by covid-19 | Carbon dioxide emissions | 5.40 | 1<br>48 | 0.02 * | 0.32 | 0.10 | $\alpha=6198$<br>$\beta<0.00001$ |
| infected by covid-19 | Nitrous oxide emissions | 5.49 | 1<br>39 | 0.02 * | 0.35 | 0.12 | $\alpha=81526$<br>$\beta=0.6683$ |
| dead by covid-19 | Nitrous oxide emissions | 2.42 | 1<br>39 | 0.12 n.s. | 0.24 | 0.058 | $\alpha=5218$<br>$\beta=0.026225$ |
| infected by covid-19 | Methane emissions | 5,73 | 1<br>39 | 0.02 * | 0.36 | 0.13 | $\alpha=82776$<br>$\beta=0.22929$ |
| dead by covid-19 | Methane emissions | 1.37 | 1<br>39 | 0.25 n.s. | 0.18 | 0.033 | $\alpha=5544$<br>$\beta=0.00672$ |
| **Social & Geographic Indicators** |  |  |  |  |  |  |  |
| infected by covid-19 | Population | 2.92 | 1<br>48 | 0.09 n.s. | 0.24 | 0.006 | $\alpha=-112655$<br>$\beta=0.00031$ |
| dead by covid-19 | Population | 0.25 | 1<br>48 | 0.61 n.s. | 0.08 | 0.006 | $\alpha=4541$<br>$\beta=130$ |
| infected by covid-19 | Area | 20,40 | 1<br>48 | 0.00004 *** | 0.55 | 0.30 | $\alpha=45752$<br>$\beta=0.05175$ |
| dead by covid-19 | Area | 11.23 | 1<br>48 | 0.001 *** | 0.43 | 0.19 | $\alpha=4051$<br>$\beta=0.0023$ |

Although Travaglio et al (2020) worked only with data from England their findings are similar to ours since they conclude that the levels of some air pollutants are linked to COVID-19 cases and morbidity^40^.

Similarly, Liu et al. (2020) studied the association of NO_2_ atmospheric level over the spread of the COVID-19 in Chinese cities, suggesting that ambient NO_2_ may contribute to the spread of COVID-19^5^. Yao et al. (2020) although studying the effect of other pollution indicators (PM_2.5_ and PM_10_) found that COVID-19 held higher death rates with increasing concentration of PM_2.5_ and PM_10_^56^.We cannot forget that pollution variables are somehow related with transport (e.g. airplanes, trains and cars)

Population and Area of the countries studied had different predictive power along the studied period. On the 21^st^ of March (C. March), both, Population and Area had high correlation with number of infected people (p=0.00007 and p=0.0043 respectively) but on the consecutive months (C. April, C. May and C. June), while Population had no statistical significance, the Area variable retained predictive power for the number of infected people. The p values for Area were p=0.009 on C. April, p=0.0002 on C. May and p=0.0004 on C. June. Similarly, during N-C May and N-C June Population variable lost its predictive power and had no significance but Area did (N-C May: p=0.00002, R^2^=0,31; N-C June: p=0.00004, R^2^=0,30).

When we considered the relation of these two variables with the number of deaths, we found similar outputs, on the 21^st^ of March (C. March) both had statistical significance (Population: p=0.014; Area: p=0.0053), but on the following cumulative months the Population variable was no longer significant while Area still was (p=0.04 on C. April; p=0.007 on C. May; p=0.0015 on C. June). Again, during N-C May and N-C June, Population loses its significance while Area retains it (N-C May: p=0,004; N-C June: p=0.001)). Aldibasi (2020) found a statistically significant positive association between Area(km^2^) and mortality rate and a negative association between Population size and incidence rate^44^ (Results are shown on Table 2 and Table 3).

### 2. Variables that are not predictive of the covid-19 pandemic (non-statistically significant linear regression p > 0.05)

None of the variables included in this group had statistical significance during the time period studied, from 21^st^ of March to the 21^st^ of June. The regression analysis with Hospital Bed Density (beds per 1000 people), Number of Physicians (per 1000 people) had no correlation with number of infected nor number of deaths at any moment in our study, either using cumulative data or non-cumulative data. When confronted with this new pandemic, health care systems showed little efficiency in preventing infections and death counts, due to probably lack of knowledge. We thought this should change with time, their predictive power had to grow as health care systems dealt with SARS-CoV-2, but astonishingly, after several months since the outbreak, all this relevant health parameters remained irrelevant. What proved to be more effective was early preventive measures. Lowental et al (2020) point in the same direction, countries that implemented early measures to limit population mixing limited the number of deaths, in this same study they explain that a delay of 7,49 days in initialling social distancing would double the number of deaths.^57^. For Papadopoulus et al (2020) early introduction of these policies are of greater importance than its stringency.^46^

Researches in R&D (per million people) or Infant Mortality Rate (indicator of a countries health and care system) showed similar behaviour. In the same line, Yao et al. (2020) did not find significance in the association between hospital beds per capita and COVID-19 death rate^56^. Sorci et al. (2020) found that the Number of hospital beds per 1,000 inhabitants was negatively associated with Case Fatality Rate ^58^. Qiu et al (2020) in their study in China, found that COVID-19 transmission is negatively moderated by the number of doctors at the city level^59^

The logic of any disease is that good health and research parameters of a country should have a positive effect over the number of infected or the number of deaths, but our results indicate no significance at all. It is necessary to highlight that the best predictor we found to death count by COVID-19 is the number of infected by COVID-19 (p>0,00001), either with cumulative data or with non-cumulative data, there is a cause-effect relation. Despite the increase in knowledge, despite improvements in health care systems… the percentage of deaths for a given number of infected could not be reduced. What it really shows is that prevention, and not treatment, is the best way to face the COVID-19 pandemic.

Unemployment rate, Inflation rate and Military expenditure, categorised as economic indicators fell also in this group of non-predictive variables. We did not find statistically significant regressions between the different economic indicators we selected and the dependent variables (number of infected and number of deaths). Richness of a country does not predict its infected or dead by COVID- 19, with the exception of those that showed easiness to travel across the country (i.e. Inbound tourism). Yao et al. among other variables they also studied Gross Domestic Product (GDP) per capita as a measurement of a country’s wealth and in their work, GDP per capita was found not to have significance in the association with COVID-19 death rate^56^. According to Qiu et al. (2020) COVID- 19 transmission is positively moderated by GDP per capita^59^

In our study, Urban population (%) and Population Growth rate were also not statistically significant in relation with number of infected people and number of deaths by SARS-CoV-2.

Government expenditure on education (as % of GDP) had some correlation with number of infected and number of deaths by SARS-CoV-2 on the 21^st^ of March but not the following two months.

The Population ages 65 and above in our study had no relation with the number of infected on the 21^st^ of March, gained significance on the 21^st^ of April (p=0.06) and went back on the 21^st^ of May to be not statistically significant. When we studied Population ages 65 and above against number of deaths it showed no correlation in March or April, but it did in May (p=0.09).

Messner also studied the effect of education and countries’ median age on the COVID-19 outbreak and found that the quality of a country’s education system is positively associated with the outbreak and that countries with an older population are more affected^60^

Shagam (2020) using multivariate linear regression found significant correlation between incidence and mortality rates with GDP per capita (p = 2.6 × 10^−15^ and 7.0 × 10^−4^, respectively), country-specific duration of the outbreak (2.6 × 10^−4^ and 0.0019), fraction of citizens over 65 years old (p = 0.0049 and 3.8 × 10^−4^) and level of press freedom (p = 0.021 and 0.019). Table 4 and table 5 can be found in complementary material.

**Table 4.** Cumulative Data. Variables that are not predictive of the covid-19 pandemic (non-statistically significant linear regression p > 0.05)

| **III: CUMULATIVE DATA. VARIABLES THAT ARE NOT PREDICTIVE OF THE COVID-19 PANDEMIC**<br>**(non-statistically significant linear regression $p > 0.05$)**<br>**21<sup>st</sup> of MARCH 2020** |  |  |  |  |  |  |  |
| --- | --- | --- | --- | --- | --- | --- | --- |
| **dependent variable (y)** | **predictive variable (x)** | **F value** | **d. f. regression residue** | **P** | **R** | **R<sup>2</sup>** | **$\alpha$ and $\beta$ coefficients** |
| **Health & Research Indicators** |  |  |  |  |  |  |  |
| infected by covid-19 | Hospital Bed Density | 0.07 | 1<br>36 | 0.78 n.s. | 0.05 | 0.00 | $\alpha=5059$<br>$\beta=-248$ |
| dead by covid-19 | Hospital Bed Density | 0.16 | 1<br>35 | 0.70 n.s. | 0.07 | 0.004 | $\alpha=380.10$<br>$\beta=-21.19$ |
| infected by covid-19 | Physicians (per 1,000 people) | 0.19 | 1<br>39 | 0.66 n.s. | 0.07 | 0.001 | $\alpha=24452$<br>$\beta=8703$ |
| dead by covid-19 | Physicians (per 1,000 people) | 0.16 | 1<br>39 | 0.69 n.s. | 0.06 | 0.004 | $\alpha=179$<br>$\beta=43.44$ |
| infected by covid-19 | Researches in R & D | 0.52 | 1<br>43 | 0.48 n.s. | 0.22 | 0.05 | $\alpha=174$<br>$\beta=4946$ |
| dead by covid-19 | Researches in R & D | 0.51 | 1<br>43 | 0.46 n.s. | 0.20 | 0.04 | $\alpha=176$<br>$\beta=4427$ |
| **Economic Indicators** |  |  |  |  |  |  |  |
| infected by covid-19 | Unemployment rate | 0.04 | 1<br>48 | 0.83 n.s. | 0.03 | 0.001 | $\alpha=4995$<br>$\beta=85$ |
| dead by covid-19 | Unemployment rate | 0.84 | 1<br>48 | 0.36 n.s. | 0.13 | 0.02 | $\alpha=96.92$<br>$\beta=21.71$ |
| infected by covid-19 | Inflation rate | 0.52 | 1<br>48 | 0.47 n.s. | 0.10 | 0.01 | $\alpha=6671$<br>$\beta=-293$ |
| dead by covid-19 | Inflation rate | 0.19 | 1<br>48 | 0.66 n.s. | 0.06 | 0.004 | $\alpha=291$<br>$\beta=-10.47$ |
| infected by covid-19 | Expenditure on education | 3.78 | 1<br>39 | 0.06 | 0.30 | 0.09 | $\alpha=21704$<br>$\beta=-3270$ |
| dead by covid-19 | Expenditure on education | 3.17 | 1<br>43 | 0.08 | 0.27 | 0.07 | $\alpha=1146$<br>$\beta=-180$ |
| infected by covid-19 | Military expenditures | 0.16 | 1<br>48 | 0.69 n.s. | 0.06 | 0.001 | $\alpha=6743$<br>$\beta=-587$ |
| dead by covid-19 | Military expenditures | 0.21 | 1<br>48 | 0.65 n.s. | 0.07 | 0.004 | $\alpha=329$<br>$\beta=-39$ |

| Social and Geographic Indicators |  |  |  |  |  |  |  |
| --- | --- | --- | --- | --- | --- | --- | --- |
| infected by covid-19 | Urban Population | 0.06 | $\frac{1}{39}$ | 0.81 n.s. | 0.04 | 0.001 | $\alpha=-8756$<br>$\beta=-34$ |
| dead by covid-19 | Urban Population | 0.07 | $\frac{1}{39}$ | 0.79 n.s. | 0.04 | 0.001 | $\alpha=-459$<br>$\beta=-2.24$ |
| infected by covid-19 | Median age | 2.80 | $\frac{1}{48}$ | 0.10 n.s. | 0.23 | 0.05 | $\alpha=-13175$<br>$\beta=503$ |
| dead by covid-19 | Median age | 1.88 | $\frac{1}{48}$ | 0.17 n.s. | 0.19 | 0.04 | $\alpha=-667$<br>$\beta=24.66$ |
| infected by covid-19 | Population ages 65 and above | 0.72 | $\frac{1}{39}$ | 0.39 n.s. | 0.13 | 0.02 | $\alpha=1959$<br>$\beta=312$ |
| dead by covid-19 | Population ages 65 and above | 1.87 | $\frac{1}{48}$ | 0.18 n.s. | 0.19 | 0.04 | $\alpha=-667$<br>$\beta=24.66$ |
| infected by covid-19 | Proportion of women in national parliaments | 0.22 | $\frac{1}{39}$ | 0.64 n.s. | 0.07 | 0.005 | $\alpha=3475$<br>$\beta=104$ |
| dead by covid-19 | Proportion of women in national parliaments | 0.21 | $\frac{1}{39}$ | 0.65 n.s. | 0.07 | 0.005 | $\alpha=-134$<br>$\beta=6.02$ |
| infected by covid-19 | Population Growth rate | 1.1 | $\frac{1}{48}$ | 0.20 n.s. | 0.15 | 0.022 | $\alpha=7756$<br>$\beta=-3411$ |
| dead by covid-19 | Population Growth rate | 0.65 | $\frac{1}{48}$ | 0.42 n.s. | 0.11 | 0.013 | $\alpha=351$<br>$\beta=-156$ |
| infected by covid-19 | Infant Mortality Rate | 0.78 | $\frac{1}{48}$ | 0.38 n.s. | 0.13 | 0.016 | $\alpha=7572$<br>$\beta=-201$ |
| dead by covid-19 | Infant Mortality Rate | 0.65 | $\frac{1}{48}$ | 0.42 n.s. | 0.12 | 0.013 | $\alpha=351$<br>$\beta=-156$ |
| infected by covid-19 | Life expectancy at birth | 1.06 | $\frac{1}{48}$ | 0.31 n.s. | 0.15 | 0.021 | $\alpha=-28178$<br>$\beta=434$ |
| dead by covid-19 | Life expectancy at birth | 0.70 | $\frac{1}{48}$ | 0.40 n.s. | 0.12 | 0.01 | $\alpha=-1388$<br>$\beta=21.12$ |
| infected by covid-19 | Obesity - Adult prevalence rate | 1.70 | $\frac{1}{39}$ | 0.20 n.s. | 0.20 | 0.004 | $\alpha=-14009$<br>$\beta=-372$ |
| dead by covid-19 | Obesity - Adult prevalence rate | 0.64 | $\frac{1}{39}$ | 0.43 n.s. | 0.13 | 0.016 | $\alpha=583$<br>$\beta=-13.80$ |

| **II: CUMULATIVE DATA. VARIABLES THAT ARE NOT PREDICTIVE OF THE COVID-19 PANDEMIC**<br>(non-statistically significant linear regression $p > 0.05$)<br>21st of APRIL 2020 | | | | | | | |
| --- | --- | --- | --- | --- | --- | --- | --- |
| dependent variable<br>(y) | predictive variable<br>(x) | F value | d. f. regression<br>residue | P | R | R<sup>2</sup> | $\alpha$ and $\beta$ coefficients |
| **Health & Research Indicators** |  |  |  |  |  |  |  |
| infected by covid-19 | Hospital Bed Density | 0.03 | 1<br>43 | 0.85 n.s. | 0.03 | 0.0006 | $\alpha=55993$<br>$\beta=-1195$ |
| dead by covid-19 | Hospital Bed Density | 0.03 | 1<br>43 | 0.84 n.s. | 0.03 | 0.0010 | $\alpha=4105$<br>$\beta=-86.81$ |
| infected by covid-19 | Physicians (per 1,000 people) | 0.44 | 1<br>43 | 0.51 n.s. | 0.10 | 0.01 | $\alpha=24452$<br>$\beta=8703$ |
| dead by covid-19 | Physicians (per 1,000 people) | 1.30 | 1<br>43 | 0.26 n.s. | 0.17 | 0.03 | $\alpha=918$<br>$\beta=961$ |
| infected by covid-19 | Researches in R & D | 0.53 | 1<br>43 | 0.47 n.s. | 0.11 | 0.01 | $\alpha=1461$<br>$\beta=5.71$ |
| dead by covid-19 | Researches in R & D | 1.74 | 1<br>43 | 0.19 n.s. | 0.19 | 0.04 | $\alpha=1313$<br>$\beta=1427$ |
| **Economic Indicators** |  |  |  |  |  |  |  |
| infected by covid-19 | Unemployment rate | 0.0002 | 1<br>43 | 0.98 n.s. | 0.002 | 0.023 | $\alpha=47418$<br>$\beta=48.05$ |
| dead by covid-19 | Unemployment rate | 0.43 | 1<br>43 | 0.43 n.s. | 0.119 | 0.014 | $\alpha=2240$<br>$\beta=176$ |
| infected by covid-19 | Inflation rate | 0.24 | 1<br>43 | 0.63 n.s. | 0.074 | 0.005 | $\alpha=53445$<br>$\beta=1768$ |
| dead by covid-19 | Inflation rate | 0.40 | 1<br>43 | 0.53 n.s. | 0.096 | 0.009 | $\alpha=3970$<br>$\beta=-147$ |
| infected by covid-19 | Expenditure on education | 0.21 | 1<br>43 | 0.65 n.s. | 0.123 | 0.015 | $\alpha=42032$<br>$\beta=-3265$ |
| dead by covid-19 | Expenditure on education | 0.0008 | 1<br>43 | 0.97 n.s. | 0.007 | 0.0001 | $\alpha=1281$<br>$\beta=10$ |
| infected by covid-19 | Military expenditures | 0.54 | 1<br>43 | 0.46 n.s. | 0.111 | 0.012 | $\alpha=30854$<br>$\beta=8962$ |
| dead by covid-19 | Military expenditures | 0.012 | 1<br>43 | 0.91 n.s. | 0.016 | 0.000 | $\alpha=3.327$<br>$\beta=87$ |

| Social & Geographic indicators |  |  |  |  |  |  |  |
| --- | --- | --- | --- | --- | --- | --- | --- |
| infected by covid-19 | Urban Population | 0.85 | $\frac{1}{43}$ | 0.36 n.s. | 0.139 | 0.019 | $\alpha=-20436$<br>$\beta=941$ |
| dead by covid-19 | Urban Population | 1.28 | $\frac{1}{43}$ | 0.26 n.s. | 0.169 | 0.028 | $\alpha=-1901$<br>$\beta=74$ |
| infected by covid-19 | Median age | 1.36 | $\frac{1}{43}$ | 0.25 n.s. | 0.175 | 0.030 | $\alpha=-64791$<br>$\beta=3004$ |
| dead by covid-19 | Median age | 2.99 | $\frac{1}{43}$ | 0.09 n.s. | 0.255 | 0.065 | $\alpha=-7165$<br>$\beta=284$ |
| infected by covid-19 | Population ages 65 and above | 1.47 | $\frac{1}{43}$ | 0.23 n.s. | 0.162 | 0.053 | $\alpha=4325$<br>$\beta=5172$ |
| dead by covid-19 | Population ages 65 and above | 3.83 | $\frac{1}{43}$ | 0.06 n.s. | 0.286 | 0.081 | $\alpha=-935$<br>$\beta=323$ |
| infected by covid-19 | Proportion of women in national parliaments | 0.241 | $\frac{1}{43}$ | 0.63 n.s. | 0.074 | 0.005 | $\alpha=26128$<br>$\beta=804$ |
| dead by covid-19 | Proportion of women in national parliaments | 2.77 | $\frac{1}{43}$ | 0.10 n.s. | 0.246 | 0.060 | $\alpha=-1128$<br>$\beta=172$ |
| infected by covid-19 | Population Growth rate | 0.21 | $\frac{1}{43}$ | 0.65 n.s. | 0.069 | 0.004 | $\alpha=56011$<br>$\beta=-12647$ |
| dead by covid-19 | Population Growth rate | 0.46 | $\frac{1}{43}$ | 0.50 n.s. | 0.103 | 0.011 | $\alpha=4286$<br>$\beta=-1220$ |
| infected by covid-19 | Infant Mortality Rate | 1.27 | $\frac{1}{43}$ | 0.26 n.s. | 0.169 | 0.028 | $\alpha=4286$<br>$\beta=-1220$ |
| dead by covid-19 | Infant Mortality Rate | 2.36 | $\frac{1}{43}$ | 0.13 n.s. | 0.228 | 0.052 | $\alpha=5284$<br>$\beta=-186$ |
| infected by covid-19 | Life expectancy at birth | 1.44 | $\frac{1}{43}$ | 0.23 n.s. | 0.179 | 0.032 | $\alpha=-282391$<br>$\beta=4237$ |
| dead by covid-19 | Life expectancy at birth | 3.31 | $\frac{1}{43}$ | 0.07 n.s. | 0.268 | 0.072 | $\alpha=-28426$<br>$\beta=409$ |
| infected by covid-19 | Obesity - Adult prevalence rate | 3.09 | $\frac{1}{43}$ | 0.09 n.s. | 0.259 | 0.007 | $\alpha=-25123$<br>$\beta=3440$ |
| dead by covid-19 | Obesity - Adult prevalence rate | 2.15 | $\frac{1}{43}$ | 0.15 n.s. | 0.218 | 0.048 | $\alpha=-506$<br>$\beta=188$ |

| **II: CUMULATIVE DATA. VARIABLES THAT ARE NOT PREDICTIVE OF THE COVID-19 PANDEMIC**<br>**(non-statistically significant linear regression $p > 0.05$)**<br>**21st of MAY 2020** |  |  |  |  |  |  |  |
| --- | --- | --- | --- | --- | --- | --- | --- |
| **dependent variable**<br>**(y)** | **predictive variable**<br>**(x)** | **F value** | **d. f. regression**<br>**residue** | **P** | **R** | **R<sup>2</sup>** | **$\alpha$ and $\beta$ coefficients** |
| **Health & Research Indicators** |  |  |  |  |  |  |  |
| infected by covid-19 | Hospital Bed Density | 0.09 | 1<br>36 | 0.76 n.s. | 0.05 | 0.0003 | $\alpha=55993$<br>$\beta=-1195$ |
| dead by covid-19 | Hospital Bed Density | 0.005 | 1<br>35 | 0.94 n.s. | 0.01 | 0.0001 | $\alpha=3755$<br>$\beta=-30.12$ |
| infected by covid-19 | Physicians (per 1,000 people) | 0.87 | 1<br>39 | 0.35 n.s. | 0.15 | 0.02 | $\alpha=35276$<br>$\beta=8492$ |
| dead by covid-19 | Physicians (per 1,000 people) | 1.77 | 1<br>39 | 0.19 n.s. | 0.21 | 0.04 | $\alpha=1084$<br>$\beta=1186$ |
| infected by covid-19 | Researches in R & D | 0.53 | 1<br>43 | 0.44 n.s. | 0.25 | 0.06 | $\alpha=71910$<br>$\beta=-779$ |
| dead by covid-19 | Researches in R & D | 0.10 | 1<br>40 | 0.19 n.s. | 0.10 | 0.009 | $\alpha=3978$<br>$\beta=-0.222$ |
| **Economic Indicators** |  |  |  |  |  |  |  |
| infected by covid-19 | Unemployment rate | 0.01 | 1<br>48 | 0.92 n.s. | 0.01 | 0.0001 | $\alpha=91570$<br>$\beta=-634$ |
| dead by covid-19 | Unemployment rate | 0.10 | 1<br>48 | 0.76 n.s. | 0.04 | 0.002 | $\alpha=5226$<br>$\beta=130$ |
| infected by covid-19 | Inflation rate | 0.13 | 1<br>43 | 0.71 n.s. | 0.05 | 0.002 | $\alpha=95252$<br>$\beta=-2281$ |
| dead by covid-19 | Inflation rate | 0.40 | 1<br>48 | 0.53 n.s. | 0.09 | 0.008 | $\alpha=7130$<br>$\beta=-266$ |
| infected by covid-19 | Expenditure on education | 0.11 | 1<br>39 | 0.74 n.s. | 0.05 | 0.002 | $\alpha=72682$<br>$\beta=-3021$ |
| dead by covid-19 | Expenditure on education | 0.0008 | 1<br>43 | 0.97 n.s. | 0.007 | 0.0001 | $\alpha=1281$<br>$\beta=10$ |
| infected by covid-19 | Military expenditures | 1.37 | 1<br>48 | 0.25 n.s. | 0.16 | 0.027 | $\alpha=37127$<br>$\beta=25781$ |
| dead by covid-19 | Military expenditures | 0.012 | 1<br>43 | 0.91 n.s. | 0.016 | 0.000 | $\alpha=3.327$<br>$\beta=87$ |

| Social & Geographic Indicators |  |  |  |  |  |  |  |
| --- | --- | --- | --- | --- | --- | --- | --- |
| infected by covid-19 | Urban Population | 0.83 | $\frac{1}{39}$ | 0.37 n.s. | 0.14 | 0.020 | $\alpha=-11100$<br>$\beta=656$ |
| dead by covid-19 | Urban Population | 1.17 | $\frac{1}{43}$ | 0.29 n.s. | 0.17 | 0.029 | $\alpha=-1233$<br>$\beta=77$ |
| infected by covid-19 | Median age | 0.36 | $\frac{1}{48}$ | 0.55 n.s. | 0.09 | 0.007 | $\alpha=-18867$<br>$\beta=2836$ |
| dead by covid-19 | Median age | 1.44 | $\frac{1}{48}$ | 0.23 ns | 0.17 | 0.03 | $\alpha=-8026$<br>$\beta=380$ |
| infected by covid-19 | Population ages 65 and above | 0.39 | $\frac{1}{39}$ | 0.39 n.s. | 0.10 | 0.010 | $\alpha=42120$<br>$\beta=1176$ |
| dead by covid-19 | Population ages 65 and above | 2.98 | $\frac{1}{39}$ | 0.09 | 0.27 | 0.071 | $\alpha=-42$<br>$\beta=314$ |
| infected by covid-19 | Proportion of women in national parliaments | <0.01 | $\frac{1}{39}$ | 0.99 n.s. | 0.00 | 0.0001 | $\alpha=58525$<br>$\beta=-2,66$ |
| dead by covid-19 | Proportion of women in national parliaments | 2.98 | $\frac{1}{39}$ | 0.09 | 0.27 | 0.071 | $\alpha=-42.20$<br>$\beta=314$ |
| infected by covid-19 | Population Growth rate | 0.06 | $\frac{1}{48}$ | 0.81 n.s. | 0.03 | 0.001 | $\alpha=94458$<br>$\beta=-12003$ |
| dead by covid-19 | Population Growth rate | 0.12 | $\frac{1}{48}$ | 0.72 n.s. | 0.05 | 0.003 | $\alpha=6925$<br>$\beta=-1207$ |
| infected by covid-19 | Infant Mortality Rate | 0.49 | $\frac{1}{48}$ | 0.49 n.s. | 0.10 | 0.010 | $\alpha=110832$<br>$\beta=-2449$ |
| dead by covid-19 | Infant Mortality Rate | 1.49 | $\frac{1}{48}$ | 0.23 n.s. | 0.17 | 0.030 | $\alpha=8956$<br>$\beta=-287$ |
| infected by covid-19 | Life expectancy at birth | 0.41 | $\frac{1}{48}$ | 0.52 n.s. | 0.09 | 0.008 | $\alpha=-236851$<br>$\beta=4166$ |
| dead by covid-19 | Life expectancy at birth | 2.27 | $\frac{1}{48}$ | 0.14 | 0.21 | 0.045 | $\alpha=-44583$<br>$\beta=652$ |
| infected by covid-19 | Obesity - Adult prevalence rate | 0.16 | $\frac{1}{39}$ | 0.68 ns | 0.06 | 0.0004 | $\alpha=45936$<br>$\beta=605$ |
| dead by covid-19 | Obesity - Adult prevalence rate | 0.13 | $\frac{1}{39}$ | 0.72 n.s. | 0.06 | 0.003 | $\alpha=3207$<br>$\beta=54$ |

| **III: CUMULATIVE DATA. VARIABLES THAT ARE NOT PREDICTIVE OF THE COVID-19 PANDEMIC**<br>**(non-statistically significant linear regression $p > 0.05$)**<br>**21<sup>st</sup> of JUNE 2020** |  |  |  |  |  |  |  |
| --- | --- | --- | --- | --- | --- | --- | --- |
| **dependent variable**<br>**(y)** | **predictive variable**<br>**(x)** | **F value** | **d. f. regression**<br>**residue** | **P** | **R** | **R<sup>2</sup>** | **$\alpha$ and $\beta$ coefficients** |
| **Health & Research Indicators** |  |  |  |  |  |  |  |
| infected by covid-19 | Hospital Bed Density | 0.38 | 1<br>36 | 0.54 n.s. | 0.10 | 0.001 | $\alpha=139284$<br>$\beta=-6798$ |
| dead by covid-19 | Hospital Bed Density | 0.39 | 1<br>35 | 0.54 n.s. | 0.10 | 0.01 | $\alpha=7600$<br>$\beta=-373$ |
| infected by covid-19 | Physicians (per 1,000 people) | 0.18 | 1<br>39 | 0.68 n.s. | 0.07 | 0.004 | $\alpha=135554$<br>$\beta=-9150$ |
| dead by covid-19 | Physicians (per 1,000 people) | 0.21 | 1<br>39 | 0.65 n.s. | 0.07 | 0.005 | $\alpha=4818$<br>$\beta=564$ |
| infected by covid-19 | Researches in R & D | 1.14 | 1<br>43 | 0.31 n.s. | 0.32 | 0.10 | $\alpha=244006$<br>$\beta=-3856$ |
| dead by covid-19 | Researches in R & D | 0.52 | 1<br>43 | 0.49 n.s. | 0.22 | 0.05 | $\alpha=10205$<br>$\beta=-1.27$ |
| **Economic Indicators** |  |  |  |  |  |  |  |
| infected by covid-19 | Unemployment rate | 0.04 | 1<br>48 | 0.84 n.s. | 0.03 | 0.001 | $\alpha=132051$<br>$\beta=1986$ |
| dead by covid-19 | Unemployment rate | 0.12 | 1<br>48 | 0.73 n.s. | 0.05 | 0.02 | $\alpha=7214$<br>$\beta=188$ |
| infected by covid-19 | Inflation rate | 0.04 | 1<br>48 | 0.84 n.s. | 0.3 | 0.0008 | $\alpha=153325$<br>$\beta=-1922$ |
| dead by covid-19 | Inflation rate | 0.21 | 1<br>48 | 0.64 n.s. | 0.07 | 0.004 | $\alpha=9496$<br>$\beta=-255$ |
| infected by covid-19 | Expenditure on education | 0.10 | 1<br>39 | 0.75 n.s. | 0.05 | 0.003 | $\alpha=77890$<br>$\beta=6940$ |
| dead by covid-19 | Expenditure on education | 0.40 | 1<br>39 | 0.53 n.s. | 0.10 | 0.01 | $\alpha=2744$<br>$\beta=767$ |
| infected by covid-19 | Military expenditures | 1.48 | 1<br>48 | 0.23 n.s. | 0.17 | 0.003 | $\alpha=60594$<br>$\beta=42273$ |
| dead by covid-19 | Military expenditures | 0.40 | 1<br>48 | 0.53 n.s. | 0.10 | 0.001 | $\alpha=2744$<br>$\beta=766$ |

| Social and Geographic Indicators |  |  |  |  |  |  |  |
| --- | --- | --- | --- | --- | --- | --- | --- |
| infected by covid-19 | Urban Population | 0.38 | $\frac{1}{39}$ | 0.54 n.s. | 0.10 | 0.001 | $\alpha=34166$<br>$\beta=1059$ |
| dead by covid-19 | Urban Population | 1.06 | $\frac{1}{39}$ | 0.31 n.s. | 0.16 | 0.003 | $\alpha=-890$<br>$\beta=100$ |
| infected by covid-19 | Median age | 0.005 | $\frac{1}{48}$ | 0.94 n.s. | 0.01 | 0.0001 | $\alpha=166722$<br>$\beta=545$ |
| dead by covid-19 | Median age | 0.37 | $\frac{1}{48}$ | 0.54 n.s. | 0.9 | 0.08 | $\alpha=-1017$<br>$\beta=257$ |
| infected by covid-19 | Population ages 65 and above | 0.77 | $\frac{1}{39}$ | 0.38 n.s. | 0.14 | 0.02 | $\alpha=164989$<br>$\beta=-3920$ |
| dead by covid-19 | Population ages 65 and above | 0.26 | $\frac{1}{39}$ | 0.61 n.s. | 0.08 | 0.007 | $\alpha=4514$<br>$\beta=131$ |
| infected by covid-19 | Proportion of women in national parliaments | 1.14 | $\frac{1}{39}$ | 0.29 n.s. | 0.17 | 0.03 | $\alpha=187929$<br>$\beta=-2851$ |
| dead by covid-19 | Proportion of women in national parliaments | 1.28 | $\frac{1}{39}$ | 0.26 n.s. | 0.18 | 0.003 | $\alpha=1686$<br>$\beta=172$ |
| infected by covid-19 | Population Growth rate | 0.002 | $\frac{1}{48}$ | 0.96 n.s. | 0.006 | 0.00004 | $\alpha=144141$<br>$\beta=3540$ |
| dead by covid-19 | Population Growth rate | 0.02 | $\frac{1}{48}$ | 0.90 n.s. | 0.02 | 0.0003 | $\alpha=8924$<br>$\beta=-561$ |
| infected by covid-19 | Infant Mortality Rate | 0.0002 | $\frac{1}{48}$ | 0.99 n.s. | 0.002 | 0 | $\alpha=147091$<br>$\beta=-74$ |
| dead by covid-19 | Infant Mortality Rate | 0.50 | $\frac{1}{48}$ | 0.48 n.s. | 0.10 | 0.01 | $\alpha=10708$<br>$\beta=-219$ |
| infected by covid-19 | Life expectancy at birth | 0.002 | $\frac{1}{48}$ | 0.96 n.s. | 0.007 | 0.00005 | $\alpha=184296$<br>$\beta=-487$ |
| dead by covid-19 | Life expectancy at birth | 0.93 | $\frac{1}{48}$ | 0.34 n.s. | 0.14 | 0.02 | $\alpha=-34683$<br>$\beta=556$ |
| infected by covid-19 | Obesity - Adult prevalence rate | 0.10 | $\frac{1}{39}$ | 0.75 n.s. | 0.05 | 0.003 | $\alpha=87007$<br>$\beta=1140$ |
| dead by covid-19 | Obesity - Adult prevalence rate | 0.19 | $\frac{1}{39}$ | 0.66 n.s. | 0.07 | 0.005 | $\alpha=5524$<br>$\beta=89$ |

**Table 5.**
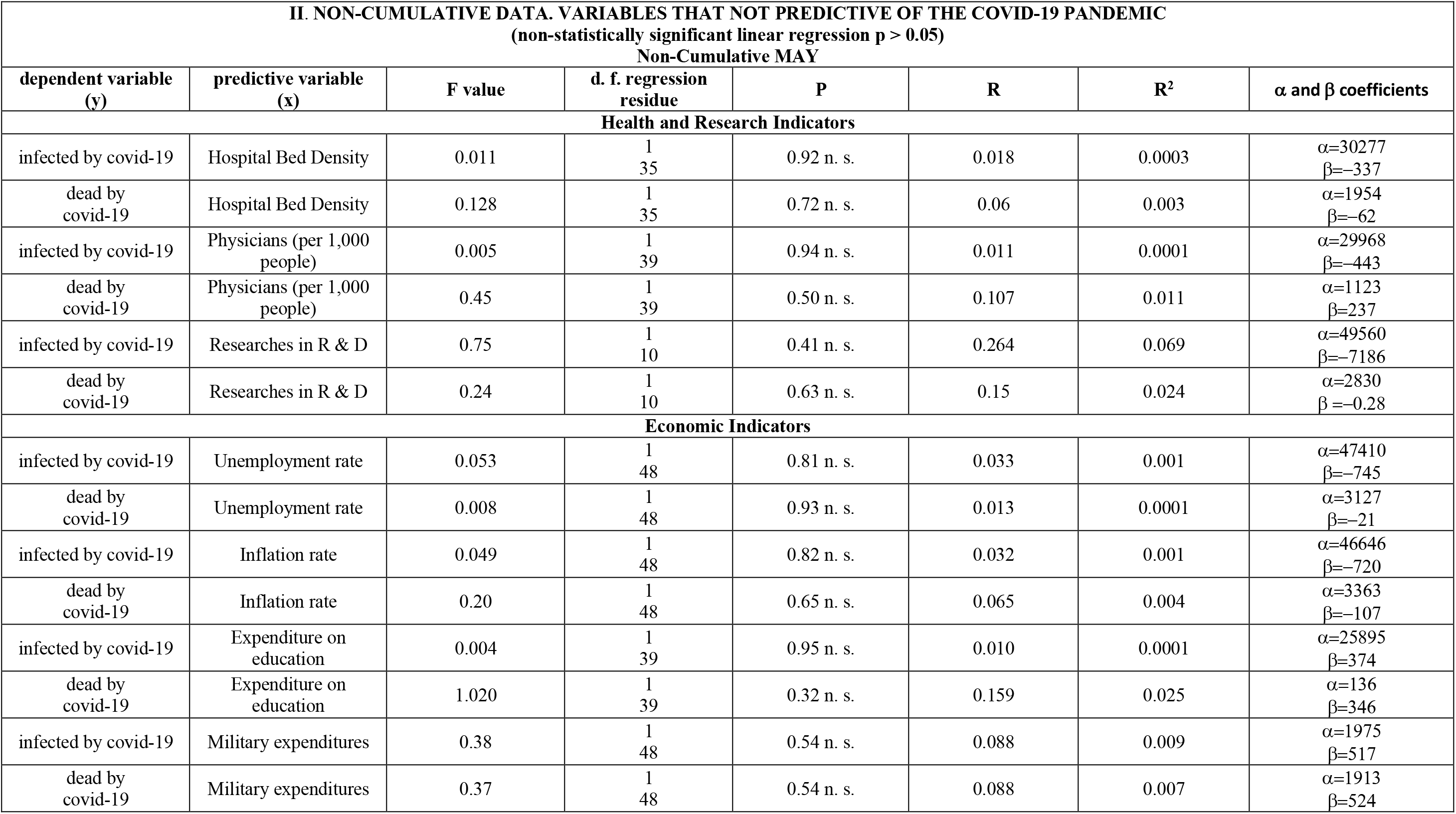
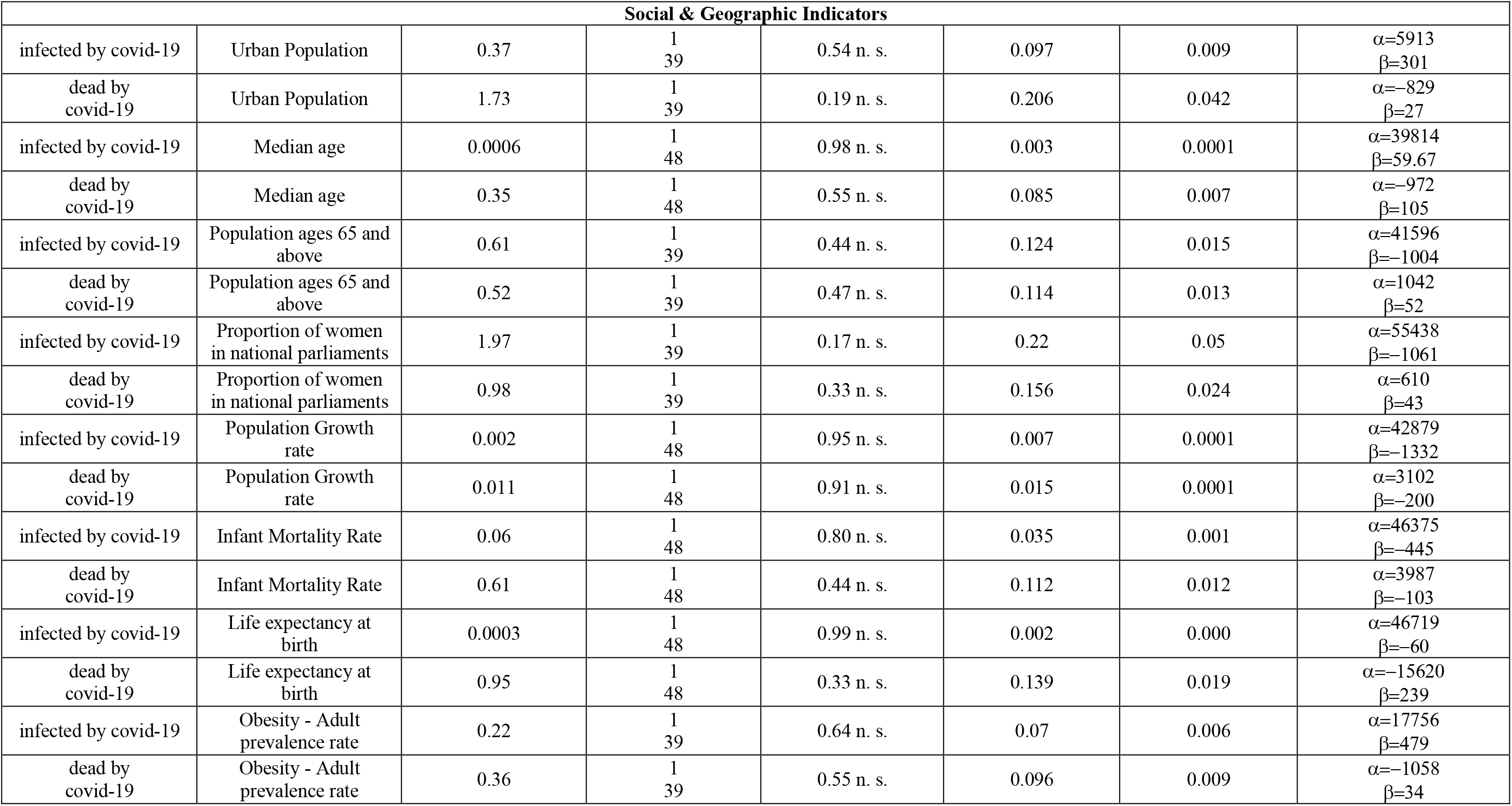
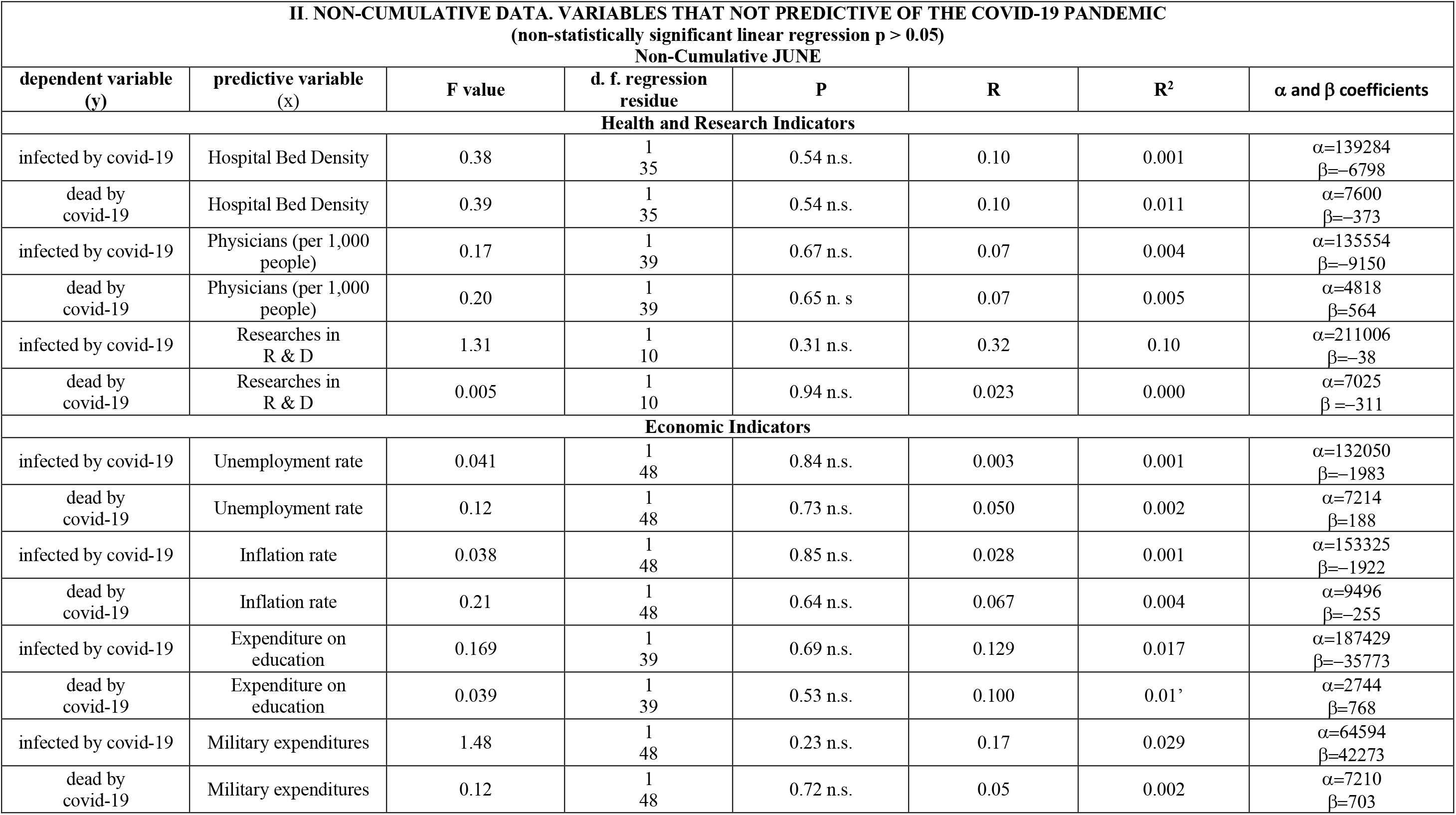
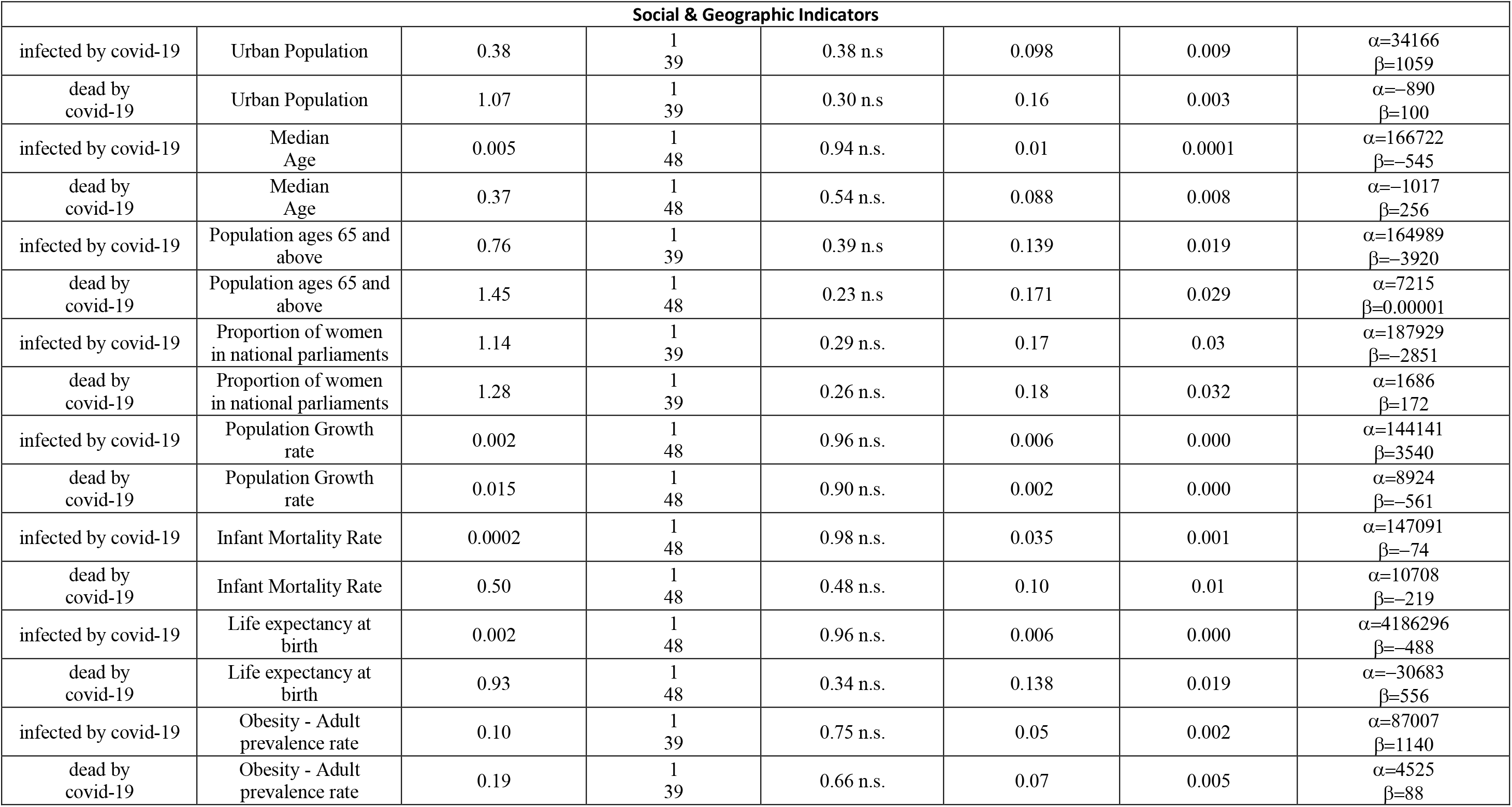
Non-Cumulative Data. Variables that are not predictive of the covid-19 pandemic (non-statistically significant linear regression p > 0.05)

Statistically, with these 50 countries (nearly 70% world’s population) number of dead depends only on number of infected according to:

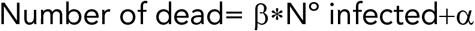

With α values ranging from -35423 to 6197 and β from 0.02910 to 16.35. The best predictor to death count by SARS-CoV2 was the number of infected (p>0,00001), either with cumulative data or with non-cumulative data, there is a cause effect relation.

The best predictive variable was inbound tourism, and it did not change throughout the time period studied, it retained its predictive strength using cumulative data or non-cumulative data.

Air pollution predictive variables are of great significance, their ability to predict number of infected and dead should make us think about the importance of One Health. Environmental health and Animal health are essential to Human health.

Despite every effort done, the increase in knowledge, the improvements implemented in health care systems, etc. yet is not enough. None of the health & research indicators had significance at all during the four-month cumulative data regressions, nor when non-cumulative data was used. As stated previously, what it really shows is that prevention, and not treatment, is the best way to confront SARS-CoV-2 outbreak, and by extension any future epidemics that may arise unknown to medicine. Some lessons must be learnt from this pandemic. Those countries that conducted more tests to isolate infectious promptly and impose to their citizens policies to prevent the spread of the virus at early stages were, by far, least affected.

According to Loewenthal et al. (2020), to reduce the number of deaths is essential to implement confinement policies at a very early stage. If social distancing is not adopted, death incidents are doubled every 7.49 days.^57^ Piguillem et al. using Italy’s early figures to calibrate their model they estimate that without lockdowns *there could be as much as 800, 000 fatalities. With a very early intervention that number reduces to about 120 – 230 fatalities, while if the intervention is started 60 days later, the number of fatalities is between 7, 200 and 9, 000*^61^. Italy had, as to 21^st^ of June, 238,275 infected people and 34,610 dead according to the World Health Organisation COVID-19 dashboard (https://covid19.who.int).

Early detection of cases through surveillance and aggressive contact tracing around known cases has helped to contain spread of the outbreak in Singapore. Together with other healthcare, border and community measures, they allow the COVID-19 outbreak to be managed without major disruption to daily living. Countries could consider these measures for a proportionate response to the risk of COVID-19^62^.

Our study shows, with very high significance, what variables predict SARS-CoV- 2 infected and dead counts In order to minimise the impact of future unknown pathogens arising, is paramount that we learn the lesson from this pandemic. Prevention is the key.

## Data Availability

All Data and links are available in the text

## ACKNOWLEDGEMENTS

We thank to Dr. C. García-Balboa, H. Díaz-Alejo, P. Martinez-Alesón for their help and advice on this paper.

## Competing Interest Statement

The authors have declared no competing interest.

## Notes

### Funding Statement

No funding was required to develop this paper

### Author Declarations

Complutense University Madrid

## BIBLIOGRAFY

1. Li, K. et al. The Clinical and Chest CT Features Associated with Severe and Critical COVID-19 Pneumonia. Invest. Radiol. (2020).

2. Zhou, G., Chen, S. & Chen, Z. Back to the spring of Wuhan: facts and hope of COVID-19 outbreak. Frontiers of Medicine (2020). doi:10.1007/s11684-020-0758-9

3. Wang, J., Tang, K., Feng, K. & Lv, W. High Temperature and High Humidity Reduce the Transmission of COVID-19. SSRN Electron. J. (2020). doi:10.2139/ssrn.3551767

4. Pirouz, B., Haghshenas, S. S., Pirouz, B., Haghshenas, S. S. & Piro, P. Development of an assessment method for investigating the impact of climate and urban parameters in confirmed cases of COVID-19: A new challenge in sustainable development. Int. J. Environ. Res. Public Health 17, (2020).

5. Liu, J. et al. Impact of meteorological factors on the COVID-19 transmission: A multi-city study in China. Sci. Total Environ. 726, 138513 (2020).

6. Guha, A., Bonsu, J., Dey, A. & Addison, D. Community and Socioeconomic Factors Associated with COVID-19 in the United States: Zip code level cross sectional analysis. medRxiv 2020.04.19.20071944 (2020). doi:10.1101/2020.04.19.20071944

7. Whittle, R. S. & Diaz-Artiles, A. An ecological study of socioeconomic predictors in detection of COVID-19 cases across neighborhoods in New York City. medRxiv 2020.04.17.20069823 (2020). doi:10.1101/2020.04.17.20069823

8. Cheng, Y. et al. Kidney impairment is associated with in-hospital death of COVID-19 patients. MedRxiv (2020).

9. Zhang, F. et al. Myocardial injury is associated with in-hospital mortality of confirmed or suspected COVID-19 in Wuhan, China: A single center retrospective cohort study. MedRxiv (2020).

10. Chen, J. et al. Clinical progression of patients with COVID-19 in Shanghai, China. J. Infect. (2020).

11. Du, R.-H. et al. Predictors of mortality for patients with COVID-19 pneumonia caused by SARS-CoV-2: a prospective cohort study. Eur. Respir. J. (2020).

12. Li, Z. et al. Caution on kidney dysfunctions of COVID-19 patients. (2020).

13. Cai, Q. et al. Experimental treatment with favipiravir for COVID-19: an open-label control study. Engineering (2020).

14. Shi, S. et al. Association of cardiac injury with mortality in hospitalized patients with COVID-19 in Wuhan, China. JAMA Cardiol. (2020).

15. Chen, C., Yan, J. T., Zhou, N., Zhao, J. P. & Wang, D. W. Analysis of myocardial injury in patients with COVID-19 and association between concomitant cardiovascular diseases and severity of COVID-19. Zhonghua Xin Xue Guan Bing Za Zhi 48, E008–E008 (2020).

16. Cheng, Y. et al. Kidney disease is associated with in-hospital death of patients with COVID-19. Kidney Int. (2020).

17. Li, J.-W. et al. The impact of 2019 novel coronavirus on heart injury: A systemic review and Meta-analysis. Prog. Cardiovasc. Dis. (2020). doi:10.1016/j.pcad.2020.04.008

18. Huang, I., Lim, M. A. & Pranata, R. Diabetes mellitus is associated with increased mortality and severity of disease in COVID-19 pneumonia – A systematic review, meta-analysis, and meta-regression: Diabetes and COVID-19. Diabetes Metab. Syndr. Clin. Res. Rev. 14, 395–403 (2020).

19. Guan, W. et al. Comorbidity and its impact on 1590 patients with Covid-19 in China: A Nationwide Analysis. Eur. Respir. J. (2020).

20. Meng, Y. et al. Sex-specific clinical characteristics and prognosis of coronavirus disease-19 infection in Wuhan, China: A retrospective study of 168 severe patients. PLOS Pathog. 16, e1008520 (2020).

21. Zhang, J. et al. Risk factors for disease severity, unimprovement, and mortality of COVID-19 patients in Wuhan, China. Clin. Microbiol. Infect. (2020). doi:10.1016/j.cmi.2020.04.012

22. Chen, R. et al. Risk factors of fatal outcome in hospitalized subjects with coronavirus disease 2019 from a nationwide analysis in China. Chest (2020). doi:10.1016/j.chest.2020.04.010

23. Zhou, F. et al. Articles Clinical course and risk factors for mortality of adult inpatients with COVID-19 in Wuhan, China: a retrospective cohort study. (2020). doi:10.1016/S0140-6736(20)30566-3

24. Aifen Lin et al. Early risk factors for the duration of SARS-CoV-2 viral positivity in COVID-19 patients. Clinical infectious diseases : an official publication of the Infectious Diseases Society of America (2020).

25. Aifen Lin et al. Early risk factors for the duration of SARS-CoV-2 viral positivity in COVID-19 patients. Clinical infectious diseases : an official publication of the Infectious Diseases Society of America (2020).

26. Du, R. H. et al. Predictors of Mortality for Patients with COVID-19 Pneumonia Caused by SARS-CoV-2: A Prospective Cohort Study. Eur. Respir. J. (2020). doi:10.1183/13993003.00524-2020

27. Yan, C. H., Faraji, F., Prajapati, D. P., Ostrander, B. T. & DeConde, A. S. Self-reported olfactory loss associates with outpatient clinical course in Covid-19. Int. Forum Allergy Rhinol. (2020). doi:10.1002/alr.22592

28. Vardavas, C. I. & Nikitara, K. COVID-19 and smoking: A systematic review of the evidence. Tob. Induc. Dis. 18, (2020).

29. Shi, P. et al. Impact of temperature on the dynamics of the COVID-19 outbreak in China. Sci. Total Environ. 728, (2020).

30. Bukhari, Q. & Jameel, Y. Will Coronavirus Pandemic Diminish by Summer? SSRN Electron. J. (2020). doi:10.2139/ssrn.3556998

31. Ficetola, G. F. & Rubolini, D. Climate affects global patterns of COVID-19 early outbreak dynamics. medRxiv 2020.03.23.20040501 (2020). doi:10.1101/2020.03.23.20040501

32. Mollalo, A., Vahedi, B. & Rivera, K. M. GIS-based spatial modeling of COVID-19 incidence rate in the continental United States. Sci. Total Environ. 728, 138884 (2020).

33. Triplett, M. Evidence that higher temperatures are associated with lower incidence of COVID-19 in pandemic state, cumulative cases reported up to March 27, 2020. medRxiv 2020.04.02.20051524 (2020). doi:10.1101/2020.04.02.20051524

34. Wang, M. et al. Temperature significant change COVID-19 Transmission in 429 cities. medRxiv 2020.02.22.20025791 (2020). doi:10.1101/2020.02.22.20025791

35. Tobías, A. & Molina, T. Is temperature reducing the transmission of COVID-19 ? Environmental Research 186, (2020).

36. Shi, P. et al. Impact of temperature on the dynamics of the COVID-19 outbreak in China. Sci. Total Environ. 138890 (2020). doi:10.1016/J.SCITOTENV.2020.138890

37. Mollalo, A., Vahedi, B. & Rivera, K. M. GIS-based spatial modeling of COVID-19 incidence rate in the continental United States. Sci. Total Environ. 138884 (2020). doi:10.1016/j.scitotenv.2020.138884

38. Luo, W. et al. The role of absolute humidity on transmission rates of the COVID-19 outbreak. (2020).

39. Hisato Takagi, Md, PhD, Toshiki Kuno, Md, PhD, Yujiro Yokoyama, M., Hiroki Ueyama, M., Takuya Matsushiro, M., Yosuke Hari, M., Tomo Ando, M. Risk and protective factors of SARS-CoV-2 infection – Meta-regression of data from worldwide nations. 1–11 (2020).

40. Travaglio, M., Popovic, R., Yu, Y., Leal, N. & Martins, L. M. Links between air pollution and COVID-19 in England. medRxiv 2020.04.16.20067405 (2020). doi:10.1101/2020.04.16.20067405

41. World Health Organisation Coronavirus Disease (COVID-19) Dashboard. Available at: https://covid19.who.int/region/euro/country/al. (Accessed: 21st April 2020)

42. World Tourism Organization. Country profile – inbound tourism | OMT.

43. Walach, H., Hockertz, S. & Sciences, M. What association do political interventions, environmental and health variables have with the number of Covid-19 cases and deaths ? (2020).

44. Aldibasi, O. S. The Association of Country-Level Factors with Outcomes of COVID-19 : Analysis of the pandemic after one million cases CURRENT STATUS : POSTED. 1–19 doi:10.21203/rs.3.rs-25834/v1

45. Daon, Y., Thompson, R. N. & Obolski, U. Estimating COVID-19 outbreak risk through air travel. medRxiv 2020.04.16.20067496 (2020). doi:10.1101/2020.04.16.20067496

46. Papadopoulos, D. I., Donkov, I., Charitopoulos, K. & Bishara, S. The impact of lockdown measures on COVID-19: a worldwide comparison. medRxiv 2020.05.22.20106476 (2020). doi:10.1101/2020.05.22.20106476

47. Chinazzi, M. et al. The effect of travel restrictions on the spread of the 2019 novel coronavirus (COVID-19) outbreak. Science (80-.). 368, 395– 400 (2020).

48. Peirlinck, M., Linka, K., Sahli Costabal, F. & Kuhl, E. Outbreak dynamics of COVID-19 in China and the United States. Biomech. Model. Mechanobiol. 1–15 (2020). doi:10.1007/s10237-020-01332-5

49. Kubota, Y., Shiono, T., Kusumoto, B. & Fujinuma, J. Multiple drivers of the COVID-19 spread: role of climate, international mobility, and region-specific conditions. medRxiv 2020.04.20.20072157 (2020). doi:10.1101/2020.04.20.20072157

50. Coelho, M. T. P. et al. Exponential phase of covid19 expansion is not driven by climate at global scale. medRxiv 2020.04.02.20050773 (2020). doi:10.1101/2020.04.02.20050773

51. Gross, B. et al. Spatio-temporal propagation of COVID-19 pandemics. 1–7 (2020).

52. Tuli, S., Tuli, S., Verma, R. & Tuli, R. Modelling for prediction of the spread and severity of COVID-19 and its association with socioeconomic factors and virus types. 1–13 (2020).

53. Koh, W. C., Naing, L. & Wong, J. Estimating the impact of physical distancing measures in containing COVID-19: an empirical analysis. medRxiv 2020.06.11.20128074 (2020). doi:10.1101/2020.06.11.20128074

54. Anzai, A., Kobayashi, T., Linton, N. M. & Kinoshita, R. Assessing the Impact of Reduced Travel on Exportation Dynamics of Novel Coronavirus Infection (COVID-19).

55. Merler, S., Pastore, A., Mu, K., Rossi, L. & Sun, K. The effect of travel restrictions on the spread of the 2019 novel coronavirus (COVID-19) outbreak. 400, 395–400 (2020).

56. Yao, Y. & Ph, D. Spatial Correlation of Particulate Matter Pollution and Death Rate of COVID-19 /China/ PM2.5,PM10/multiple linear regression. (2020).

57. Loewenthal, G. et al. COVID-19 pandemic-related lockdown: response time is more important than its strictness. medRxiv 2020.06.11.20128520 (2020). doi:10.1101/2020.06.11.20128520

58. Sorci, G., Faivre, B. & Morand, S. Why does COVID-19 case fatality rate vary among countries? medRxiv 2020.04.17.20069393 (2020). doi:10.1101/2020.04.17.20069393

59. Qiu, Y., Chen, X. & Shi, W. Impacts of social and economic factors on the transmission of coronavirus disease 2019 (COVID-19) in China. J. Popul. Econ. 2019, 1–27 (2020).

60. Messner, W. The Institutional and Cultural Context of Cross-National Variation in COVID-19 Outbreaks. medRxiv 2019, 2020.03.30.20047589 (2020).

61. Piguillem, F. The Optimal COVID-19 Quarantine and Testing. (2020).

62. Lee, V. J., Chiew, C. J. & Khong, W. X. Interrupting transmission of COVID-19 : lessons from containment efforts in Singapore. 1–5 (2020). doi:10.1093/jtm/taaa039

